# Vaccine-Hesitant Parents’ Considerations Regarding Covid-19 Vaccination of Adolescents

**DOI:** 10.1101/2021.05.25.21257780

**Authors:** Erga Atad, Itamar Netzer, Orr Peleg, Keren Landsman, Keren Dalyot, Shanny Edan Reuven, Ayelet Baram-Tsabari

## Abstract

**Introduction:** Israel led a rapid vaccine rollout against COVID-19, leading to a local remission of the epidemic and rolling back of most public health measures. Further vaccination of 12-15-year-olds may be hindered by public perceptions of the necessity and safety of vaccination.

**Methods:** we examined the considerations of vaccine hesitant parents (VHPs) regarding vaccination of children against COVID-19. The responses of 456 parents were surveyed and analyzed before FDA authorization of vaccination of children.

**Results:** parents who were vaccinated against COVID-19 were more likely to intend to vaccinate their children (r=-0.466, p<0.01). Low accessibility of vaccination may be a dissuading factor for VHPs more inclined to vaccinate. Vaccine efficacy and gaining a “Green Pass” were positively associated with an intention to vaccinate and statistically significant. VHPs inclined not to vaccinate indicated short development time and possible long term effects as dissuading factors.

**Discussion:** vaccine promotion should be tailored for VHP’s positive and negative considerations for higher uptake.

## Introduction

Israel has led a rapid vaccine rollout to combat COVID-19, starting with vaccination of medical personnel, high risk populations and senior citizens^1^. This resulted in more than five million citizens receiving both Pfizer-Biontech COVID-19 vaccination doses, out of 9.2 million (5,092,562, 54.60%^2^.) The success of a vaccination campaign for 12-15-year-olds, another 6.7% of the population^3^, rests on communicating the necessity, safety and efficacy of the vaccine to them and their legal guardians. This is further challenged by the current remission of the epidemic, the rolling back of most public health measures in Israel, the low morbidity associated with COVID-19 infection in children^4^, and the rapid rollout of a novel vaccine under an FDA Emergency Use Authorization.

We examined the considerations of vaccine hesitant parents (VHPs) about COVID-19 vaccinations for their children. This is a pressing need as the FDA approved the immediate vaccination of 12-15-year-olds on May 10th 2021^5^, and a vaccination campaign in Israel is expected to commence shortly. Previous studies have found that vaccine hesitancy is associated with inaccurate knowledge, beliefs and perceptions on vaccines, negative attitudes and behaviors toward vaccination, and demographic characteristics (many or older children, low levels of income and education)^6–8^

## Methods

Institutional review board approval was received from the Technion - Israel Institute of Technology.

We conducted a digital survey using a commercial online data collection provider (iPanel, Bnei Brak, Israel). A sample of 1118 Hebrew-speaking Israelis who are parents of 12-15-year-old children consented to complete a survey before the vaccine was approved (28 April to 6 May, 2021). A link was provided to a Qualtrics survey. Total compensation did not exceed 20 NIS (6 US dollars). The survey was based on earlier COVID-19 related surveys ^6^. Respondents were first asked to answer a 5-point Likert screening question: “Do you intend to vaccinate your children when the COVID-19 vaccine becomes available for them?”. We analyzed the responses of parents who met minimum requirements (completion of the entire survey including an open-ended question, a minimum of 3 minutes spent answering, and correct response to an attention question, n=456).

## Results

Overall, parents were very positive regarding their intention to vaccinate their 12-15 children against COVID-19. Among the 1118 parents 31.5% answered they will definitely vaccinate their children, 26.2% “will probably vaccinate”, 27.4% were neutral, 8.8% chose “probably will not vaccinate”, and 6.3% “will definitely not vaccinate”. Parents responding they are likely to vaccinate made 57.8% of the respondents.

We analyzed the responses of 456 parents, including 38 who answered “1 – Definitely will not vaccinate”, 50 who answered “5 – Definitely will vaccinate”, (capped from a total of 352) and 368 parents who responded 2, 3 or 4 on the screening question and we define as being VHPs. Among this sample, parents who were vaccinated against COVID-19 were more likely to intend to vaccinate their children (r=-0.466, p<0.01). Respondents with family income above average were more inclined to vaccinate than those with average income or below (F=(2, 441)=5.98, p<0.05).

The main considerations for and against vaccination are presented in Table 1. Among these considerations, fear of COVID-19 variants, side effects of the vaccine and possible long-term effects were most listed as affecting the decision to vaccinate. Speed of vaccine development was a more typical concern of parents who reported lower intention to vaccinate, as well as unique health characteristics of one’s child (not significant). Low accessibility of vaccination may be a dissuading factor for VHPs more inclined to vaccinate. Vaccine efficacy, gaining a “Green Pass”^9^ and social responsibility (not significant) were positively associated with an intention to vaccinate. Factor analysis demonstrates differing sets of considerations for or against vaccination for parents inclined to provide or withhold vaccinations.

**Table 1:**
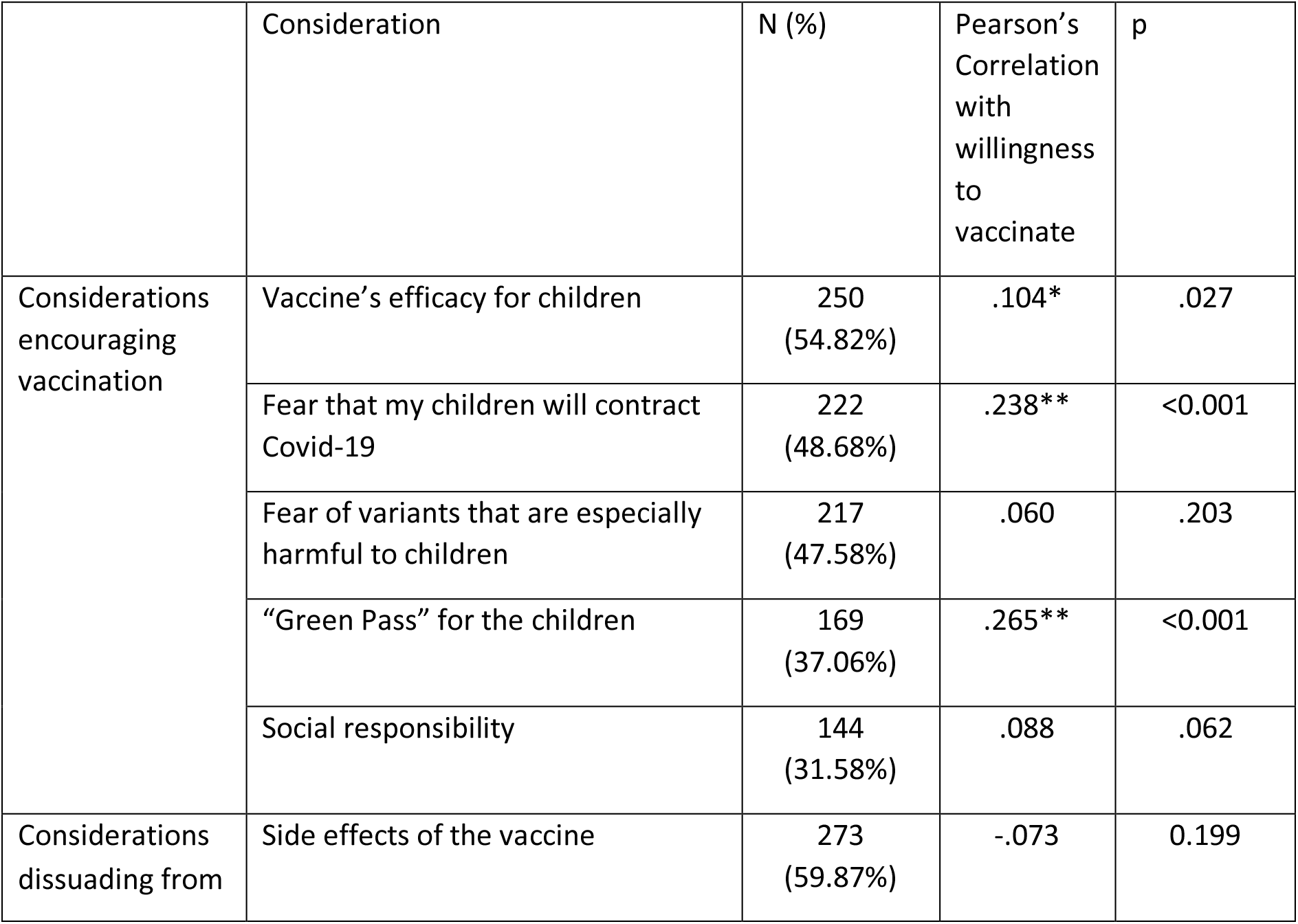
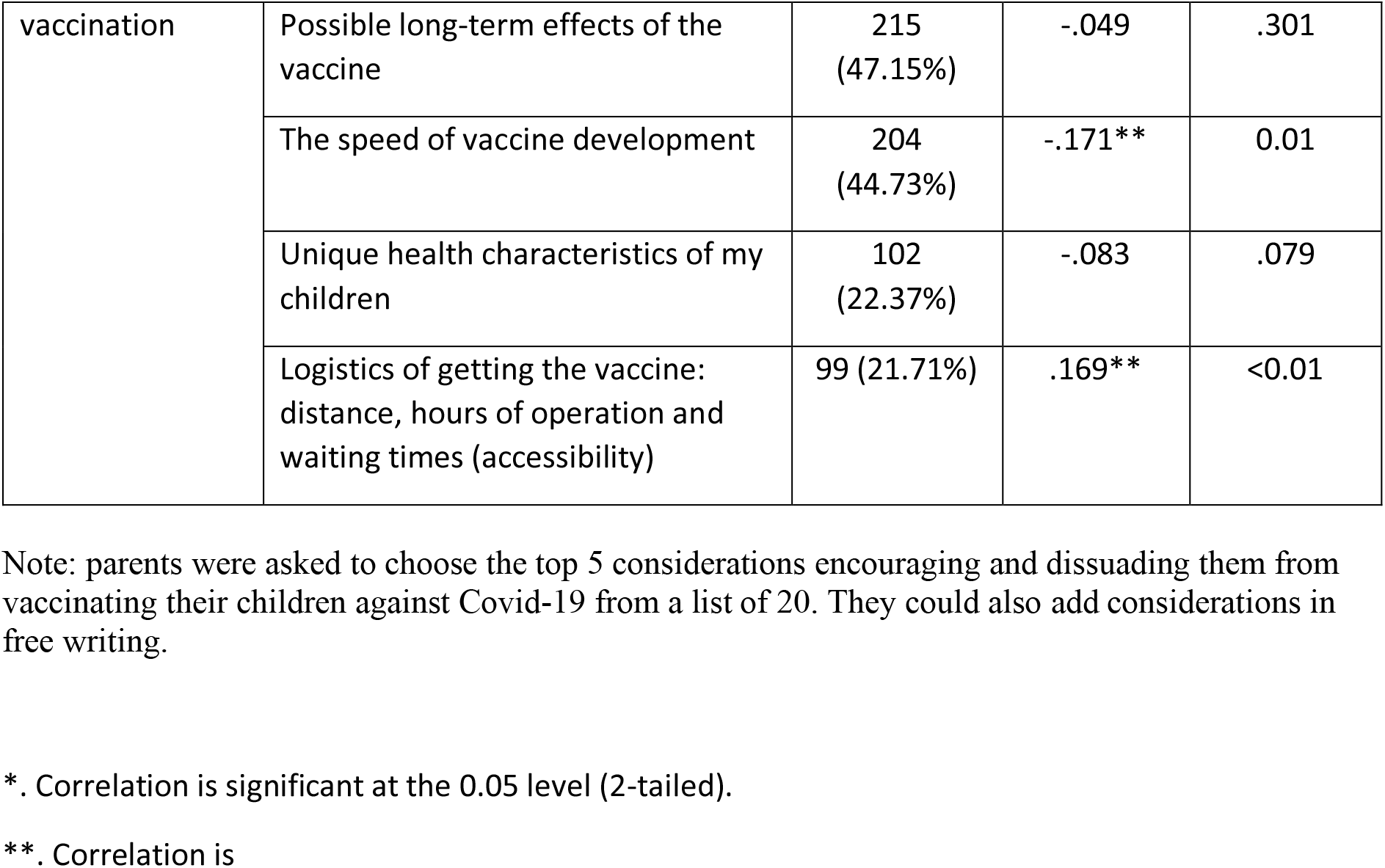
Considerations encouraging and dissuading vaccination and their correlation with willingness to vaccinate.

## Discussion

As the coming phase of the vaccination campaign targets adolescents who are at a lower risk from COVID-19, public health officials should address theirs and their legal guardians’ concerns, whether legitimate or misinformed^10^. These initial findings indicate that considerations for and against vaccination differ among VHPs inclined to vaccinate or not vaccinate their children. Campaign messages should be tailored to reinforce encouraging considerations and address discouraging ones, to assist in attaining high vaccine uptake.

The second stage of this study will conduct a paired follow-up questionnaire several weeks into a campaign to vaccinate 12–15-year-olds, to discover how VHPs decided to act and what impacted their decision to administer or withhold vaccines from their children.

## Data Availability

All raw data is available upon request

